# Elevated Cardiac Biomarkers in Relatively Healthy U.S. Adults

**DOI:** 10.1101/2023.11.27.23299072

**Authors:** Sophie E. Claudel, Insa M. Schmidt, Deepa M. Gopal, Ashish Verma

**Affiliations:** Department of Medicine, Boston Chobanian & Avedisian School of Medicine, Boston, MA; Renal Section, Department of Medicine, Boston University Chobanian & Avedisian School of Medicine, Boston, MA; Section of Cardiology, Department of Medicine, Boston University Chobanian & Avedisian School of Medicine, Boston, MA

## Abstract

**Background:** Despite the importance of early cardiovascular disease (CVD) intervention, little data exists for evaluating cardiovascular risk in adults without traditional CVD risk factors (e.g., diabetes, hypertension).

**Methods:** We included 4,544 adults from the 1999-2004 National Health and Nutrition Examination Survey without prevalent diabetes, hypertension, chronic kidney disease, or CVD. We used multi-variable adjusted Cox proportional hazards regression modeling to assess the relationship between logarithmically transformed cardiac biomarkers (high sensitivity cardiac troponin T (hs-cTnT), hs-cTnI (Abbott, Ortho, and Siemens assays), and NT-proBNP) and CVD mortality among a nationally representative cohort of relatively healthy adults.

**Results:** The mean age was 38.2 years (SD 12.8) and 53.9% were women. 8.7% had elevated levels of hs-cTnT or NT-proBNP above previously established thresholds for subclinical CVD. In multivariable adjusted models, each doubling of hs-cTnT was associated with a 49% increased risk of CVD mortality (Hazard Ratio (HR) 1.49, 95%CI 1.02-2.17, *p*=0.04). Only two of the hs-cTnI assays (Abbott and Ortho) were significantly associated with CVD mortality (Abbott HR 1.48, 95%CI 1.06-2.07, *p*=0.02; Ortho HR 1.47, 95%CI 1.23-1.77, *p*=0.0001). Each doubling of NT-proBNP was associated with a 41% increased risk of CVD mortality (HR 1.38, 95%CI 1.09-1.74, *p*=0.008).

**Conclusion:** Younger patients who maintain relatively good health may still carry occult CVD risk. Efforts to reduce population-wide CVD should consider novel methods for risk stratification, as standard CVD risk factors may overlook subpopulations at risk.

**Clinical Significance:**

- Among relatively healthy adults without significant CVD risk factors, elevated cardiac troponins and NT-proBNP are associated with cardiovascular and all-cause mortality.
- Subclinical cardiovascular disease poses higher cardiovascular risk among relatively healthy adults than previously appreciated.
- Efforts to reduce population-wide CVD should consider novel methods for risk stratification and early intervention to mitigate preventable CVD.

## Introduction

Cardiovascular disease (CVD) is the leading cause of death in the United States.^1^ Prevention of CVD events often relies on early identification of higher risk individuals based on known CVD risk factors. However, subclinical CVD risk may go undetected in individuals without traditional risk factors such as diabetes, hypertension, hyperlipidemia, and obesity.^1^ Novel approaches to cardiovascular risk stratification in younger, healthier adults are needed to inform clinical care and CVD prevention.^2^ Whether adults with serologic evidence of subclinical CVD, but without traditional CVD risk factors, are at increased risk of adverse cardiovascular outcomes is not well understood. As a result, there are no current guidelines for managing CVD risk in this population.

We assessed the relationship between subclinical CVD as determined by cardiac biomarkers (high-sensitivity cardiac troponins T and I (hs-cTnT, hs-cTnI) and N-terminal-pro-brain natriuretic peptide (NT-proBNP)) and mortality among healthy adults participating in the National Health and Nutrition Examination Survey (NHANES).

## Methods

We included 4,544 adults ages 20 years and older who participated in NHANES 1999-2004. We excluded those with prevalent CVD (coronary artery disease, myocardial infarction, angina, stroke, heart failure), diabetes, hypertension, estimated glomerular filtration rate (eGFR) <60 ml/min/1.73m^2^, urinary albumin-to-creatinine ratio (UACR) ≥30 mg/g, and those who underwent dialysis treatment within the last year. Prevalent CVD was determined by participant self-report. Diabetes was defined by hemoglobin A1c ≥6.4%, self-report, or use of hypoglycemic medications. Hypertension was defined as a blood pressure ≥130/80 mmHg, self-report, or use of antihypertensive medications. In a sensitivity analysis, we further excluded patients with hyperlipidemia (defined by use of a statin or total cholesterol ≥240 mg/dL) and active smoking (N=2,524).

### Cardiac biomarkers and subclinical CVD

In NHANES, hs-cTnT was measured using a Roche Cobas e601 (Elecsys reagents), which has a lower limit of detection (LLOD) of 3 ng/L. hs-cTnI (Abbott) was measured using an Abbott ARCHITECT i2000SR and had a LLOD of 1.7 ng/L. hs-cTnI (Ortho) was measured using an Ortho Vitros 36000 and had a LLOD of 0.39 ng/L. hs-cTnI (Siemens) was measured using a Siemens Centaur XP and had a lower limit of detection of 1.6 ng/L. NT-proBNP was measured using a Roche Cobas e601 autoanalyzer (LLOD 5 pg/ml, upper LOD 35,000 pg/ml). Details including coefficients of variability for each assay are publicly available at https://wwwn.cdc.gov/Nchs/Nhanes/1999-2000/SSTROP_A.htm. Only samples that had not previously undergone a freeze-thaw cycle were included in the analyses.

Subclinical CVD was defined using thresholds established in the literature: NT-proBNP ≥125 pg/mL, hs-cTnT ≥14 ng/L among women, or hs-cTnT ≥22 ng/L among men.^3^ In a sensitivity analysis, subclinical CVD was separately defined by hs-cTnI (Abbott) ≥10 ng/L in women or ≥12 ng/L in men.^4^

### Outcome ascertainment

The mortality status of participants was ascertained through linkage with the National Center for Health Statistics National Death Index through December 31^st^, 2019. Cardiovascular mortality included death from heart disease (ICD-10 codes I00-I09, I11, I13, I20-I151) and cerebrovascular diseases (ICD-10 codes I60-I69).

### Statistical Analysis

Baseline demographic and clinical characteristics were assessed for the overall population and stratified by presence of subclinical CVD. Multivariable-adjusted Cox proportional hazards models were used to explore the association between cardiac biomarkers and all-cause mortality and CVD mortality. Models were controlled for age, sex, race or ethnicity, health insurance status, smoking status, survey year, food insecurity, body mass index, hemoglobin A1c, total cholesterol, systolic blood pressure, insufficient physical activity, use of a statin, use of an angiotensin converting enzyme inhibitor (ACEi) or angiotensin receptor blocker (ARB), and eGFR.

We performed several sensitivity analyses. First, we excluded individuals who reported active smoking and had hyperlipidemia (defined by use of a statin or total cholesterol ≥240 mg/dL). We performed the original analyses stratified by the presence of subclinical CVD. Finally, we assessed the association between subclinical CVD (as a categorical variable) and mortality.

All analyses were performed with sample weights according to NHANES analytic guidelines. All analyses were performed using SAS version 9.4 (SAS Institutes, Cary, North Carolina). A two-sided *p*-value <0.05 was considered statistically significant.

## Results

The mean (standard deviation) age was 38.2 (12.8) years, 53.9% of the sample were women and 71.4% were non-Hispanic White (**Table 1**). The median hs-cTnT was 3.4 ng/L [interquartile range (IQR) 2.6, 4.3] among women and 5.3 ng/L [IQR 4.1, 6.8] among men and the median NT-proBNP was 37.7 pg/mL [IQR 20.0, 68.2]. The median hs-cTnI (Abbott) was 1.26 ng/L [0.81, 1.91], the median hs-cTnI (Ortho) was 0.21 ng/L [0, 0.50], and the median hs-cTnI (Siemens) was 1.18 ng/L [IQR 0.44, 2.52]. Overall crude prevalence of subclinical CVD was 8.7% in the whole population. The prevalence was higher in women (13.0%) compared to men (3.7%). The median follow-up time was 17.6 years and there were 83 CVD mortality events and 441 all-cause mortality events overall. Among those with subclinical CVD, there were 33 CVD mortality events and 138 all-cause mortality events.

**Table 1.** Baseline characteristics of the population.

|  | **Overall**<br>**(N=4,544)** | **No Subclinical CVD**<br>**(N=4,136)** | **Subclinical CVD <sup>a</sup>**<br>**(N=408)** |
| --- | --- | --- | --- |
| Age (years) | 38.2 (12.8) | 37.1 (13.6) | 49.3 (16.0) |
| Female | 53.9 | 51.4 | 80.5 |
| Race or ethnicity |  |  |  |
| Non-Hispanic White | 71.4 | 70.1 | 84.2 |
| Mexican American | 9.2 | 9.7 | 3.7 |
| Non-Hispanic Black | 8.5 | 8.9 | 4.7 |
| Other | 10.9 | 11.3 | 7.4 |
| Poverty-to-income ratio (PIR) | 3.1 [1.6, 5.0] | 3.1 [1.6, 5.0] | 3.2 [1.7, 5.0] |
| Income |  |  |  |
| Low Income (PIR <1) | 13.6 | 13.8 | 11.9 |
| Middle Income (PIR 1-3) | 35.0 | 35.0 | 35.0 |
| High Income (PIR >3) | 51.4 | 51.2 | 53.1 |
| Education |  |  |  |
| Below High School | 16.5 | 16.5 | 15.7 |
| High School/GED | 24.1 | 24.2 | 22.7 |
| Above High School | 59.5 | 59.2 | 61.6 |
| Uninsured | 22.6 | 23.5 | 12.8 |
| Food insecure | 11.0 | 11.3 | 7.2 |
| Current/former smoking | 48.0 | 47.5 | 53.0 |
| Body mass index (kg/m<sup>2</sup>) | 26.6 (5.5) | 26.7 (6.1) | 26.1 (6.5) |
| Hemoglobin A1c (%) | 5.2 (0.3) | 5.2 (0.8) | 5.2 (1.0) |
| LDL cholesterol (mg/dL) | 117.7 (23.0) | 118.2 (23.4) | 111.8 (23.0) |
| HDL cholesterol (mg/dL) | 52.9 (15.1) | 52.4 (15.1) | 58.0 (17.1) |
| Total cholesterol (mg/dL) | 195.4 (39.7) | 195.3 (41.0) | 195.5 (40.1) |
| eGFR (ml/min per 1.73m<sup>2</sup>) | 102.8 (16.0) | 103.6 (16.3) | 94.4 (16.1) |
| UACR (mg/g) | 4.9 [3.5, 7.5] | 4.8 [3.5, 7.3] | 5.6 [3.9, 8.7] |
| Statin use | 2.4 | 2.2 | 4.5 |
| hsTnT (ng/L) | 4.1 [3.2, 5.7] | 4.1 [3.2, 5.6] | 4.7 [3.4, 8.2] |
| hsTnI-Abbott (ng/L) | 1.3 [0.8, 1.9] | 1.3 [0.8, 1.9] | 1.3 [0.8, 2.3] |
| hsTnI-Ortho (ng/L) | 0.2 [0, 0.5] | 0.2 [0, 0.5] | 0.3 [0.05, 0.8] |
| hsTnI-Siemens (ng/L) | 1.8 [0.8, 3.4] | 1.8 [0.8, 3.3] | 2.0 [0.8, 4.1] |
| NT-proBNP (pg/mL) | 37.1 [20.0, 68.2] | 34.0 [18.5, 59.2] | 160.0 [138.8, 211.4] |
| Subclinical CVD |  |  |  |
| Defined by hs-cTnT <sup>b</sup> | 1.0 | 0 | 11.9 |
| Defined by hs-cTnI (Abbott) <sup>c</sup> | 1.1 | 0.8 | 3.8 |
| Defined by NT-proBNP <sup>d</sup> | 8.0 | 0 | 92.0 |

<sup>a</sup> Defined by either elevated hs-cTnT <sup>b</sup> or elevated NT-proBNP <sup>d</sup> <sup>b</sup> hs-cTnT: ≥14 ng/L among men and ≥22 ng/L among women <sup>c</sup> hs-cTnI (Abbott): ≥12 ng/L among men and ≥10 ng/L among women <sup>d</sup> NT-proBNP: ≥125 pg/mL

In multivariable-adjusted models, each doubling of hs-cTnT was associated with a 49% increased risk of cardiovascular mortality (hazard ratio (HR) 1.49, 95% confidence interval (CI) 1.02-2.17, *p*=0.04) and a 37% increased risk of all-cause mortality (HR 1.37, 95% CI 1.12-1.68, *p*=0.003) (**Table 2**). Both the Abbott and Ortho hs-cTnI assays were significantly associated with both cardiovascular and all-cause mortality. In contrast, hs-cTnI Siemens assay was not significantly associated with either CVD mortality (HR 1.1, 95% CI 0.80-1.63, *p*=0.47) or all-cause mortality (HR 1.09, 95% CI 0.97-1.22, *p*=0.16) after adjusting for multiple clinical and demographic variables. Each doubling of NT-proBNP was associated with a 41% increased risk of cardiovascular mortality (HR 1.38, 95% CI 1.09-1.74, *p*=0.008) and a 23% increased risk of all-cause mortality (HR 1.23, 95% CI 1.12-1.34, *p*<0.0001).

**Table 2.**
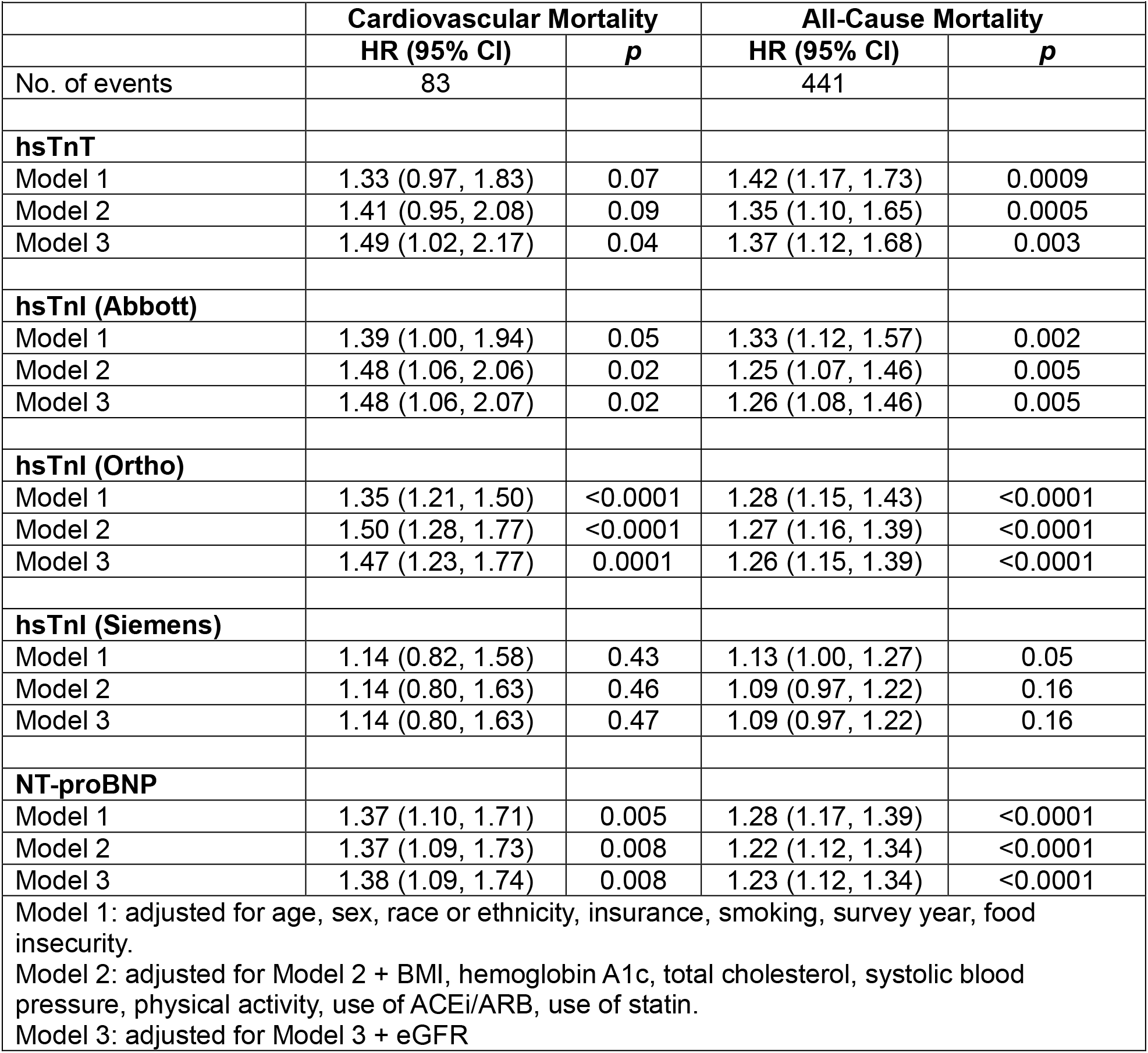
The association of cardiac biomarkers with cardiovascular and all-cause mortality.

|  |  |  |  |  |
| --- | --- | --- | --- | --- |
|  | **Cardiovascular Mortality** |  | **All-Cause Mortality** |  |
|  | **HR (95% CI)** | **p** | **HR (95% CI)** | **p** |
| No. of events | 83 |  | 441 |  |
| **hsTnT** |  |  |  |  |
| Model 1 | 1.33 (0.97, 1.83) | 0.07 | 1.42 (1.17, 1.73) | 0.0009 |
| Model 2 | 1.41 (0.95, 2.08) | 0.09 | 1.35 (1.10, 1.65) | 0.0005 |
| Model 3 | 1.49 (1.02, 2.17) | 0.04 | 1.37 (1.12, 1.68) | 0.003 |
| **hsTnI (Abbott)** |  |  |  |  |
| Model 1 | 1.39 (1.00, 1.94) | 0.05 | 1.33 (1.12, 1.57) | 0.002 |
| Model 2 | 1.48 (1.06, 2.06) | 0.02 | 1.25 (1.07, 1.46) | 0.005 |
| Model 3 | 1.48 (1.06, 2.07) | 0.02 | 1.26 (1.08, 1.46) | 0.005 |
| **hsTnI (Ortho)** |  |  |  |  |
| Model 1 | 1.35 (1.21, 1.50) | <0.0001 | 1.28 (1.15, 1.43) | <0.0001 |
| Model 2 | 1.50 (1.28, 1.77) | <0.0001 | 1.27 (1.16, 1.39) | <0.0001 |
| Model 3 | 1.47 (1.23, 1.77) | 0.0001 | 1.26 (1.15, 1.39) | <0.0001 |
| **hsTnI (Siemens)** |  |  |  |  |
| Model 1 | 1.14 (0.82, 1.58) | 0.43 | 1.13 (1.00, 1.27) | 0.05 |
| Model 2 | 1.14 (0.80, 1.63) | 0.46 | 1.09 (0.97, 1.22) | 0.16 |
| Model 3 | 1.14 (0.80, 1.63) | 0.47 | 1.09 (0.97, 1.22) | 0.16 |
| **NT-proBNP** |  |  |  |  |
| Model 1 | 1.37 (1.10, 1.71) | 0.005 | 1.28 (1.17, 1.39) | <0.0001 |
| Model 2 | 1.37 (1.09, 1.73) | 0.008 | 1.22 (1.12, 1.34) | <0.0001 |
| Model 3 | 1.38 (1.09, 1.74) | 0.008 | 1.23 (1.12, 1.34) | <0.0001 |
| Model 1: adjusted for age, sex, race or ethnicity, insurance, smoking, survey year, food insecurity. |  |  |  |  |
| Model 2: adjusted for Model 2 + BMI, hemoglobin A1c, total cholesterol, systolic blood pressure, physical activity, use of ACEi/ARB, use of statin. |  |  |  |  |
| Model 3: adjusted for Model 3 + eGFR |  |  |  |  |

A sensitivity analysis excluding patients with existing hyperlipidemia and current smoking showed similar findings, with larger effect sizes for the associations between cardiac biomarkers and mortality in some cases (**Table S1**). Stratifying by subclinical CVD showed attenuation of the relationship between each biomarker and all-cause mortality to non-significance among those without subclinical CVD (**Table S2**). Subclinical CVD, as a dichotomous variable, was not significantly associated with CVD mortality when defined by hs-cTnT and NT-proBNP or by hs-cTnI alone (**Table S3**).

## Discussion

In this sample of asymptomatic, community-dwelling adults without known CVD, chronic kidney disease, diabetes, or hypertension, nearly 9% had evidence of subclinical CVD. Both hs-cTnT and NT-proBNP were significantly associated with CVD and all-cause mortality. This finding expands prior work showing an association between these biomarkers and mortality among higher risk patients with CVD risk factors (e.g., diabetes, hypertension).^3,5–7^

The heterogeneity in the association between high-sensitivity cardiac troponin assays and mortality based on assay used suggests that this biomarker may not be suitable for screening for subclinical CVD in the general population. Indeed, prior literature has suggested that hs-cTnI may represent an entirely different pathophysiologic mechanism than hs-cTnT; where hs-cTnT is associated with cardiac structural changes, hs-cTnI may represent atherosclerotic burden.^8^

Traditional CVD risk factors include diabetes, hypertension, hyperlipidemia, chronic kidney disease, obesity, and smoking and patients are counseled and treated to prevent or control these conditions. However, our findings show even younger patients who maintain good health may still carry occult CVD risk. Monitoring traditional, large macro-level risk factors may be insufficient for risk stratification, and further advances in precision medicine may be needed to identify CVD risk in younger populations.^9^ This finding challenges the conventional approach to identifying and screening higher risk adults, at which point disease may be well-established and more difficult or expensive to treat.^2^ The lack of an association after excluding participants with subclinical CVD suggests that the clinical thresholds for defining subclinical CVD are reasonable. By showing the risk associated with subclinical CVD in a population without traditional CVD risk factors, our findings highlight the need for preventative measures to reduce cardiovascular morbidity and mortality at the population level.^10^

This analysis is limited by one time measurement of cardiac biomarkers. Additionally, the ascertainment of all-cause and CVD death in NHANES was based exclusively on ICD codes. A large proportion of patients were actively smoking at the time of their NHANES exam and while the analyses were controlled for smoking status, residual confounding from other lifestyle factors may still be present. Nonetheless, a sensitivity analysis excluding patients with hyperlipidemia and active smoking demonstrated similar results. There are several additional strengths to this manuscript, including the use of a nationally-representative, community dwelling population of asymptomatic adults who would not normally be screened for CVD in clinical settings, and inclusion of covariates that capture socio-demographic determinants of health (food insecurity, health insurance, physical activity).^11^

In summary, subclinical cardiovascular disease poses higher cardiovascular risk among apparently healthy adults than previously appreciated. Efforts to reduce population-wide CVD should consider novel methods for risk stratification and early intervention to mitigate preventable CVD.

## Data availability statement

Data are available in a repository and can be accessed via a DOI link (https://wwwn.cdc.gov/nchs/nhanes/Default.aspx).

## References

1. Tsao CW, Aday AW, Almarzooq ZI, et al. Heart Disease and Stroke Statistics - 2023 Update: A Report from the American Heart Association. Vol 147.; 2023. doi:10.1161/CIR.0000000000001123

2. Turco JV, Inal-Veith A, Fuster V. Cardiovascular Health Promotion: An Issue That Can No Longer Wait. Journal of the American College of Cardiology. 2018;72(8):908–913. doi:10.1016/j.jacc.2018.07.007

3. Fang M, Wang D, Tang O, et al. Subclinical Cardiovascular Disease in US Adults With and Without Diabetes. Journal of the American Heart Association. 2023;12. doi:10.1161/JAHA.122.029083

4. Abbott Laboratories. Abbott’s High Sensitive Troponin-I. Published 2018. Accessed April 10, 2023. https://www.corelaboratory.abbott/int/en/offerings/assays/risk-stratification

5. Hillis GS, Welsh P, Chalmers J, et al. The relative and combined ability of high-sensitivity cardiac troponin t and n-terminal pro-b-type natriuretic peptide to predict cardiovascular events and death in patients with type 2 diabetes. Diabetes Care. 2014;37(1):295–303. doi:10.2337/dc13-1165

6. McEvoy JW, Daya N, Tang O, et al. High-sensitivity troponins and mortality in the general population. Eur Heart J. 2023;353(2023):1–11. doi:10.1093/eurheartj/ehad328

7. Saunders JT, Nambi V, De Lemos JA, et al. Cardiac troponin T measured by a highly sensitive assay predicts coronary heart disease, heart failure, and mortality in the atherosclerosis risk in communities study. Circulation. 2011;123(13):1367–1376. doi:10.1161/CIRCULATIONAHA.110.005264

8. Lemos JA De, Berry JD. Comparisons of multiple troponin assays for detecting chronic myocardial injury in the general population: redundant or complementary ? Eur Heart J. 2023;00:1–3.

9. Leopold JA, Loscalzo J. Emerging role of precision medicine in cardiovascular disease. Circulation Research. 2018;122(9):1302–1315. doi:10.1161/CIRCRESAHA.117.310782

10. Vaduganathan M, Mensah GA, Turco JV, Fuster V, Roth GA. The Global Burden of Cardiovascular Diseases and Risk: A Compass for Future Health. Journal of the American College of Cardiology. 2022;80(25):2361–2371. doi:10.1016/j.jacc.2022.11.005

11. Powell-Wiley TM, Baumer Y, Baah FO, et al. Social Determinants of Cardiovascular Disease. Circulation Research. 2022;130(5):782–799. doi:10.1161/CIRCRESAHA.121.319811

