## Supplemental Tables for "Elevated Cardiac Biomarkers in Relatively Healthy U.S. Adults"

### SUPPLEMENTARY MATERIALS

**Table S1. The association of cardiac biomarkers with cardiovascular and all-cause mortality, excluding participants with hyperlipidemia and current smoking.**

| | Cardiovascular Mortality | | All-Cause Mortality | |
|---|---|---|---|---|
| | **HR (95% CI)** | ***p*** | **HR (95% CI)** | ***p*** |
| No. of events | 43 | | 185 | |
| **hsTnT** | 4.84 (1.26, 18.46) | 0.02 | 1.64 (1.28, 2.09) | 0.0002 |
| **hsTnI (Abbott)** | 2.04 (1.41, 2.96) | 0.0004 | 1.39 (1.14, 1.68) | 0.002 |
| **hsTnI (Ortho)** | 1.52 (1.21, 1.92) | 0.0007 | 1.00 (0.35, 2.85) | 1.00 |
| **hsTnI (Siemens)** | 1.31 (0.84, 2.04) | 0.22 | 1.20 (1.01, 1.44) | 0.04 |
| **NT-proBNP** | 2.25 (1.16, 4.33) | 0.02 | 1.23 (1.09, 1.39) | 0.002 |

Modes adjusted for age, sex, race or ethnicity, insurance, former smoking, survey year, food insecurity, BMI, hemoglobin A1c, total cholesterol, systolic blood pressure, physical activity, use of ACEi/ARB, use of statin, eGFR

**Table S2. The association of cardiac biomarkers with all-cause mortality among adults without subclinical CVD.**

| | Without Subclinical CVD | | With Subclinical CVD | |
|---|---|---|---|---|
| | **HR (95% CI)** | ***p*** | **HR (95% CI)** | ***p*** |
| No. of events | 303 | | 138 | |
| **hsTnT** | | | | |
| Model 1 | 1.26 (0.93, 1.71) | 0.13 | 1.39 (1.03, 1.86) | 0.03 |
| Model 2 | 1.17 (0.87, 1.58) | 0.29 | 1.45 (1.05, 1.99) | 0.02 |
| Model 3 | 1.20 (0.88, 1.64) | 0.25 | 1.47 (1.07, 2.02) | 0.02 |
| **hsTnI (Abbott)** | | | | |
| Model 1 | 1.10 (0.89, 1.37) | 0.37 | 1.70 (1.24, 2.32) | 0.001 |
| Model 2 | 1.08 (0.86, 1.35) | 0.52 | 1.55 (1.12, 2.14) | 0.009 |
| Model 3 | 1.08 (0.86, 1.35) | 0.50 | 1.55 (1.12, 2.14) | 0.009 |
| **hsTnI (Ortho)** | | | | |
| Model 1 | 1.13 (0.97, 1.32) | 0.11 | 1.84 (1.43, 2.37) | <0.0001 |
| Model 2 | 1.13 (0.94, 1.37) | 0.19 | 1.68 (1.30, 2.17) | 0.0002 |
| Model 3 | 1.13 (0.94, 1.36) | 0.19 | 1.68 (1.30, 2.17) | 0.0002 |
| **hsTnI (Siemens)** | | | | |
| Model 1 | 1.06 (0.92, 1.23) | 0.42 | 1.22 (0.95, 1.56) | 0.11 |
| Model 2 | 1.05 (0.90, 1.21) | 0.55 | 1.12 (0.87, 1.44) | 0.37 |
| Model 3 | 1.05 (0.90, 1.20) | 0.54 | 1.12 (0.87, 1.44) | 0.37 |
| **NT-proBNP** | | | | |
| Model 1 | 1.16 (1.01, 1.33) | 0.04 | 1.34 (1.12, 1.61) | 0.002 |
| Model 2 | 1.13 (0.98, 1.30) | 0.08 | 1.31 (1.06, 1.66) | 0.03 |
| Model 3 | 1.14 (0.99, 1.31) | 0.08 | 1.31 (1.03, 1.67) | 0.03 |

| |
|---|
| Model 1: adjusted for age, sex, race or ethnicity, insurance, smoking, survey year, food insecurity.<br>Model 2: adjusted for Model 2 + BMI, hemoglobin A1c, total cholesterol, systolic blood pressure, physical activity, use of ACEi/ARB, use of statin.<br>Model 3: adjusted for Model 3 + eGFR |

**Table S3. The association of subclinical CVD with cardiovascular and all-cause mortality.**

| | Cardiovascular Mortality | | All-Cause Mortality | |
|---|---|---|---|---|
| | **HR (95% CI)** | ***p*** | **HR (95% CI)** | ***p*** |
| No. of events | 83 | | 441 | |
| **Subclinical CVD defined by hs-cTnT and NT-proBNP [a]** | | | | |
| Crude | 6.75 (3.58, 12.73) | <0.0001 | 4.23 (2.37, 5.49) | <0.0001 |
| Model 1 | 2.15 (1.03, 4.51) | 0.04 | 1.87 (1.33, 1.62) | 0.0006 |
| Model 2 | 2.00 (0.82, 4.88) | 0.13 | 1.62 (1.09, 2.42) | 0.02 |
| Model 3 | 1.97 (0.78, 4.97) | 0.15 | 1.62 (1.08, 2.42) | 0.02 |
| **Subclinical CVD defined by hs-cTnI (Abbott) [b]** | | | | |
| Crude | 3.00 (0.36, 24.17) | 0.30 | 2.57 (1.08, 6.12) | 0.03 |
| Model 1 | 2.37 (0.36, 15.76) | 0.36 | 2.24 (1.17, 4.27) | 0.02 |
| Model 2 | 2.29 (0.27, 19.39) | 0.44 | 1.81 (0.89, 3.69) | 0.10 |
| Model 3 | 2.27 (0.26, 19.70) | 0.45 | 1.81 (0.88, 3.69) | 0.10 |

Model 1: adjusted for age, sex, race or ethnicity, insurance, smoking status, survey year, food insecurity.
Model 2: adjusted for Model 2 + BMI, hemoglobin A1c, total cholesterol, systolic blood pressure, physical activity, use of ACEi/ARB, use of statin.
Model 3: adjusted for Model 3 + eGFR

[a] Subclinical CVD defined by hs-cTnT ≥22 ng/L among men or hs-cTnT ≥14 ng/L among women or NT-proBNP >125 pg/mL.
[b] Subclinical CVD defined by hs-cTnI (Abbott) ≥12 ng/L among men and ≥10 ng/L among women.
